# Antibiotic receipt by respiratory symptom group and source of care among children under five in Bangladesh: repeated cross-sectional analyses of national surveys, 2019 and 2025

**DOI:** 10.64898/2026.11.11.26367825

**Authors:** S. M. Ashraf Ul Hoque, Md Khairul Islam, Mohammad Khalid Hasan, Tanvir Ahmed, Ashika Sultana, S M Raysul Haque, Fariha Haseen, Kamran ul Baset

## Abstract

**Background:** Antibiotic use for childhood respiratory illness is common in low-income and middle-income countries, but national surveys rarely separate symptom groups or care pathways. We estimated antibiotic receipt among children under five in Bangladesh by reported respiratory symptom group, source of care and household wealth.

**Methods:** We analysed the 2019 and 2025 Bangladesh Multiple Indicator Cluster Surveys. We included 4,010 children with cough and difficult breathing in the previous two weeks (3,990 in regression models). We compared breathing difficulty from the chest (suspected pneumonia) with nose-only symptoms, and five reported sources of care. Survey-weighted modified Poisson regression estimated adjusted prevalence ratios (PRs). We described where antibiotic-treated children sought care and estimated slope indices of inequality (SIIs) by wealth.

**Results:** In 2025, 68.5% (95% CI 64.2 to 72.5) of children with suspected pneumonia received an antibiotic, as did 54.3% (51.3 to 57.2) of children with nose-only symptoms. In pooled analyses of both surveys, the adjusted PR for nose-only symptoms was 0.83 (0.78 to 0.89). Against public health care, the pooled adjusted PR was 0.98 (0.91 to 1.07) for pharmacy care without a medical facility. It was 0.24 (0.20 to 0.29) for no advice or treatment sought. Pharmacy care rose from 18.8% to 36.5% of children with cough and difficult breathing between 2019 and 2025. By 2025, it accounted for 43.7% of treated children with suspected pneumonia and 42.2% with nose-only symptoms. Across the pooled surveys, receipt for suspected pneumonia was higher in richer children (SII 12.5 percentage points, 1.0 to 24.0). Recorded amoxicillin receipt in suspected pneumonia was 17.7% (14.7 to 21.1) in 2025, with no clear wealth gradient.

**Conclusions:** Antibiotic receipt was common in both symptom groups, and care contacts shifted towards pharmacies. Receipt and care seeking differed by wealth. Antibiotic receipt should be monitored alongside symptoms and care contacts. Stewardship should be evaluated in medical and pharmacy settings while protecting access to assessment and appropriate treatment. The surveys cannot determine whether individual courses were appropriate.

## Background

Antimicrobial resistance was associated with almost 4.7 million deaths in 2021, and the burden is forecast to grow [1]. Antibiotic consumption has risen fastest in low-income and middle-income countries (LMICs) [2]. Among sick children under five, reported antibiotic use across national surveys from 73 countries rose from 36.8% in 2005 to 43.1% in 2017 [3]. Children in these countries receive an estimated 18.5 antibiotic courses before age five, much of it for illnesses that do not need antibiotics [4 to 6]. Yet some children do need them. For instance, the World Health Organization (WHO) recommends oral amoxicillin, an Access antibiotic, for children aged 2 to 59 months with fast breathing or chest indrawing pneumonia [7,8]. By contrast, antibiotics do not help the common cold or a runny nose [9]. Stewardship therefore has to reduce unnecessary use without limiting treatment for children who need it.

Bangladesh shows this tension clearly. Here, sick children are taken to public facilities, private and non-governmental (NGO) providers, pharmacies and informal providers [10]. The National Drug Policy prohibits the sale of antibiotics without a prescription [11], and the Drugs and Cosmetics Act 2023 added a penalty for such sales [12]. The national antimicrobial resistance strategy for 2023 to 2028 also plans to restrict over-the-counter sales and to introduce stewardship in pharmacies [13]. In practice, drug shops sell antibiotics over the counter, and village doctors (rural practitioners without formal medical qualifications) and other informal providers prescribe them widely [11,14,15]. The choice of drug also matters. More than half of the antibiotics available in Bangladesh are in the WHO Watch group, which carries a higher risk of resistance [8,16].

To guide stewardship, policy makers need to know which children receive antibiotics and where they seek care. However, national evidence answers these questions only in part. Earlier studies described antibiotic receipt for respiratory symptoms or pooled childhood illnesses, but did not report the chest-related versus nose-only symptom contrast examined here [17 to 20]. Caregiver and hospital studies found heavy over-the-counter use but were not nationally representative [10,21]. Multi-country studies, in turn, showed rising use and the role of the source of care, without separating symptom groups [3,4,22,23]. Household surveys cannot diagnose pneumonia [24,25]. They do, however, ask whether difficult breathing came from the chest or a blocked nose, which allows an operational comparison of two symptom groups. Pharmacies already supply many antibiotics [11,14,15], but we found no national analysis that links pharmacy care to antibiotic receipt in recent surveys.

To address these gaps, we analysed the 2019 and 2025 Bangladesh Multiple Indicator Cluster Surveys (MICS). We estimated antibiotic receipt among children under five with reported cough and difficult breathing, comparing chest-related and nose-only symptoms. We then examined associations with reported sources of care, changes in care seeking and the distribution of antibiotic-treated children, and wealth-related inequalities. Recorded antibiotic types and the distinction between advice sources and medicine acquisition were examined as secondary analyses.

## Methods

### Data sources and participants

MICS are nationally representative household surveys run by the Bangladesh Bureau of Statistics with support from the United Nations Children’s Fund (UNICEF) [26]. Each round used a two-stage stratified cluster sample [27 to 30]. Census enumeration areas were selected within strata, and 20 households were then drawn systematically in each selected area. In each sampled household, mothers or caretakers answered an under-five questionnaire for every child under five. The response rate for this questionnaire was 93.6% in 2019 and 94.6% in 2025. Fieldwork ran from January to June 2019 and from February to June 2025. Trained teams of interviewers led by a supervisor collected the data on tablets, and supervisors re-interviewed one household per cluster.

We included all children with a completed under-five questionnaire, identified by a positive under-five sample weight. The earlier 2006 and 2012-13 rounds were used only to check the published antibiotic figures. The analysis followed the Strengthening the Reporting of Observational Studies in Epidemiology (STROBE) guidance for cross-sectional studies [31]. Supplementary tables and figures are in Additional file 1.

### Symptom groups

Caregivers reported illness in the two weeks before the interview. They were first asked about cough and about fast or difficult breathing. If the child had difficult breathing, they were asked whether it came from a problem in the chest, a blocked or runny nose, or both. We required both cough and difficult breathing, because earlier rounds asked the breathing question only of children with cough (Table S1).

From these reports we defined two operational groups. Suspected pneumonia, the MICS definition used before 2025, was cough with difficult breathing from the chest only or from both chest and nose. The 2025 report dropped the cough requirement, and we reproduced its figure separately (Table S2). By contrast, the nose-only group had cough with difficult breathing from a blocked or runny nose only. Pneumonia is more likely in the first group, but clinical need is uncertain in both, and concurrent illnesses may also prompt treatment [24,25]. The groups therefore cannot show whether an individual antibiotic course was appropriate.

### Antibiotic receipt

Caregivers were asked whether the child was given any medicine for the illness at any time, and then named all medicines given. Interviewers then coded them (Table S1). In 2019 the question was asked for fever or cough, and in 2025 for difficult breathing, so every child in the study population was asked. Antibiotics were coded as amoxicillin, cotrimoxazole, other antibiotic pill or syrup, and other antibiotic injection or intravenous (IV). Separately, one code recorded medicines known only by brand name and another recorded medicines of unknown type. Neither was counted as an antibiotic in the primary analysis.

### Source of care

Caregivers listed every place where they sought advice or treatment. We assigned one source per child, giving priority to the most formal medical contact: public health care, then private or NGO medical source, then pharmacy, then informal provider. Under this rule, children with a pharmacy contact but no public or private medical contact formed the pharmacy group. We call this pharmacy care in the text and label it “pharmacy, no medical facility” in tables and figures. Informal providers were traditional practitioners, relatives, shops and other places. The last group sought no advice or treatment, which does not mean that no medicine was given.

Because one source was assigned per child, some groups include other contacts. In the pharmacy group, 17 children in 2019 and 8 in 2025 also reported an informal provider, and higher-priority groups could also include lower-level contacts (Table S3). The source of care records where advice was sought, not who prescribed or supplied the antibiotic, and MICS has no separate code for village doctors. In 2019 only, caregivers also reported where the antibiotic was obtained. For brevity, we refer to the first three groups together as “a public, private or pharmacy source”, which does not imply contact with a clinician.

### Statistical analysis

All estimates used the child sample weights, clusters and strata. Symptom groups were analysed as subpopulations, so all children contributed to variance estimation. Variances used Taylor linearisation with survey-specific strata (Table S4). The MICS weights are normalised within each survey, so pooled estimates weight 2019 and 2025 by sample size. Percentages are weighted and counts are unweighted. Proportions have logit-based 95% confidence intervals (CIs). We first reproduced the antibiotic figures in the four survey reports (Table S2). This agreement supports consistency with the published coding, but it does not validate how medicines were classified.

#### Antibiotic receipt by symptom group

We estimated receipt in each symptom group and survey. Differences between surveys and between groups are percentage point differences from survey-weighted linear models. We also estimated the share of antibiotic-treated children who had nose-only symptoms.

#### Associations with symptom group and source of care

We pooled the two surveys and used survey-weighted modified Poisson regression to estimate prevalence ratios (PRs) [32]. Each exposure had its own model. The first compared nose-only symptoms with suspected pneumonia. The second compared five sources of care, with public health care as the reference. We chose confounders a priori from earlier work on antibiotic use in children and from what both surveys measured [3,17,18,22]. They were survey, child age (0 to 11, 12 to 23, 24 to 35, 36 to 59 months) and sex. They also included diarrhoea in the same two weeks, mother’s education, wealth quintile, residence and division (eight administrative regions). MICS builds the wealth index from household assets and dwelling characteristics with principal component analysis [29,30]. The source model also adjusted for symptom location, because symptoms influence both where a child is taken and whether an antibiotic is given. Fever could not be included, because the 2025 survey did not ask about it.

We report crude, minimally adjusted (survey, child age and sex) and fully adjusted PRs, and adjusted prevalence differences averaged over the weighted model sample. A few fitted probabilities in the source model exceeded 1, so we checked the differences with logistic standardisation, which gave similar results (Table S4). However, source of care probably lies partly on the path from symptoms to antibiotic use, because symptoms may influence whether and where a child is taken. We therefore present the symptom model without source as the main estimate. Adding source compares children with the same type of care; it does not give a direct effect. We also tested each exposure by survey interaction. Finally, models used complete cases without imputation. They excluded children with breathing difficulty from another or unknown location or with missing data (Table S3).

#### Changes in care seeking and the distribution of treated children

We described the source of care in each survey. Post hoc, we described how antibiotic-treated children were distributed across the five sources. We also estimated, post hoc, the percentage of all children with cough and difficult breathing who both received an antibiotic and had each source (Table S5). These measures show where observed receipt is concentrated, not how likely receipt is within a source or how much use was inappropriate. A Kitagawa decomposition then divided the 2019 to 2025 change in receipt into parts due to where children went and to receipt within each source [33]. The decomposition is descriptive, and its CIs came from the delta method.

#### Wealth inequalities

We estimated the slope index of inequality (SII) with survey-weighted linear models on ridit-scored wealth [34]. Ridit scores were the midpoints of the weighted cumulative wealth quintile shares within each symptom group and survey. Pooled models adjusted for survey and assumed a common slope in both surveys. The SII is the modelled percentage point gap between the top and bottom of the wealth distribution.

#### Secondary analyses

We grouped antibiotic types among treated children into five mutually exclusive patterns. These were any injection or IV, amoxicillin only, amoxicillin with another oral antibiotic, cotrimoxazole without amoxicillin, and other oral antibiotic only. Post hoc, we estimated the SII for amoxicillin, and amoxicillin receipt in children aged 2 to 59 months, the age range of WHO pneumonia guidance [7]. We also cross-tabulated the 2019 source of care against where the antibiotic was obtained.

#### Sensitivity analyses

Each analysis changed one assumption (Table S6). For the outcome, we counted brand-name-only medicines as antibiotics. In a separate unknown-type reclassification scenario, we counted medicines of unknown type as antibiotics. This scenario did not reclassify other or brand-name-only codes, so it is not an exhaustive upper bound on antibiotic receipt. Other analyses used logistic regression, added interview season (winter, summer, monsoon, post-monsoon) or fitted each survey separately. For symptom location, we also excluded children with diarrhoea and used chest-only symptoms as the reference. For source of care, we also combined informal care with no care. Post hoc, we added fever to the 2019 models and excluded the 25 pharmacy-group children who also reported an informal provider (Tables S4 and S5). To gauge unmeasured confounding, for example by illness severity or fever, we computed E-values [35].

We report estimates with 95% CIs and two-sided p values, without a significance threshold. Analyses used Stata version 17 (StataCorp, College Station, Texas, USA), and figures used R version 4.6.1 (R Foundation for Statistical Computing, Vienna, Austria).

#### Use of large language models

The authors used Claude Sonnet 5.5 (Anthropic) for grammar and language editing, improving readability and flow, and formatting. The authors reviewed all edits and take full responsibility for the manuscript.

## Results

In 2019 and 2025, 1,829 and 2,181 children had cough with difficult breathing, 4,010 in total (Table S3). Of these, 468 and 637 had suspected pneumonia (Table 2). After 20 exclusions for another or unknown symptom location or missing data, 3,990 children entered the regression models (Table S3). About three in four children in the model sample lived in rural areas in both surveys (Table 1). Our estimates also matched the published antibiotic figures for suspected pneumonia in all four survey rounds (Table S2).

**Table 1.** Characteristics of children with cough and difficult breathing in the model sample, Bangladesh MICS 2019 and 2025, n (weighted %).

| Characteristic | 2019, n (weighted %) | 2025, n (weighted %) |
|---|---|---|
| All children in the model sample | 1,827 (100) | 2,163 (100) |
| **Symptoms** |  |  |
| Chest only | 300 (14.4) | 333 (15.8) |
| Chest and nose | 168 (9.8) | 304 (14.2) |
| Nose only | 1,359 (75.8) | 1,526 (70.0) |
| **Child age** |  |  |
| 0 to 11 months | 488 (26.5) | 545 (25.1) |
| 12 to 23 months | 431 (24.1) | 578 (27.9) |
| 24 to 35 months | 382 (21.1) | 428 (18.7) |
| 36 to 59 months | 526 (28.4) | 612 (28.3) |
| **Sex** |  |  |
| Boy | 1,032 (55.9) | 1,187 (54.7) |
| Girl | 795 (44.1) | 976 (45.3) |
| **Diarrhoea in the same two weeks** |  |  |
| No | 1,597 (87.5) | 1,911 (88.2) |
| Yes | 230 (12.5) | 252 (11.8) |
| **Mother's education** |  |  |
| None or pre-primary | 168 (9.7) | 125 (5.4) |
| Primary | 453 (24.0) | 387 (17.9) |
| Secondary | 912 (48.4) | 1,213 (55.3) |
| Higher secondary or above | 294 (17.9) | 438 (21.5) |
| **Wealth quintile** |  |  |
| Lowest (Quintile 1 / Poorest) | 476 (22.5) | 533 (21.4) |
| Second (Quintile 2 / Poorer) | 426 (21.9) | 476 (21.3) |
| Middle (Quintile 3) | 298 (16.1) | 423 (19.7) |
| Fourth (Quintile 4 / Richer) | 338 (18.8) | 421 (21.2) |
| Highest (Quintile 5 / Richest) | 289 (20.7) | 310 (16.4) |
| **Residence** |  |  |
| Rural | 1,464 (76.7) | 1,616 (73.0) |
| Urban | 363 (23.3) | 547 (27.0) |
| **Division** |  |  |
| Barishal | 194 (6.5) | 207 (6.6) |
| Chattogram | 378 (22.4) | 651 (32.0) |
| Dhaka | 240 (16.3) | 263 (13.2) |
| Khulna | 297 (12.1) | 202 (6.9) |
| Mymensingh | 210 (14.5) | 106 (6.6) |
| Rajshahi | 234 (14.8) | 253 (14.2) |
| Rangpur | 248 (12.2) | 346 (14.8) |
| Sylhet | 26 (1.3) | 135 (5.6) |
| **Source of care** |  |  |
| Public health care | 264 (14.4) | 260 (11.5) |
| Private or NGO medical | 378 (20.3) | 386 (18.0) |
| Pharmacy, no medical facility | 324 (18.8) | 787 (36.6) |
| Informal provider | 460 (24.0) | 366 (18.1) |
| No advice or treatment sought | 401 (22.4) | 364 (15.7) |

**Table 2.** Antibiotic receipt and related care by respiratory symptom group, Bangladesh MICS 2019 and 2025, % (95% CI).

| Measure | 2019 | 2025 |
|---|---|---|
| **Chest-related symptoms: suspected pneumonia** |  |  |
| Antibiotic given | 62.9 (57.7 to 67.8); n = 468 | 68.5 (64.2 to 72.5); n = 637 |
| Chest only | 61.5 (55.2 to 67.4); n = 300 | 64.7 (58.2 to 70.7); n = 333 |
| Chest and nose | 65.0 (56.4 to 72.7); n = 168 | 72.7 (66.8 to 77.9); n = 304 |
| Care from a public, private or pharmacy source | 68.2 (63.3 to 72.7); n = 468 | 77.5 (73.6 to 81.0); n = 637 |
| Amoxicillin given | 16.6 (13.0 to 20.9); n = 468 | 17.7 (14.7 to 21.1); n = 637 |
| **Nose-only symptoms: cough with difficult breathing from the nose only** |  |  |
| Share of children with cough and difficult breathing | 75.8 (73.6 to 77.9); n = 1,829 | 69.3 (67.0 to 71.5); n = 2,181 |
| Antibiotic given | 54.0 (51.1 to 57.0); n = 1,360 | 54.3 (51.3 to 57.2); n = 1,526 |
| Care from a public, private or pharmacy source | 48.9 (45.7 to 52.0); n = 1,360 | 61.3 (58.2 to 64.2); n = 1,526 |
| **All children with cough and difficult breathing** |  |  |
| Antibiotic given | 56.2 (53.6 to 58.7); n = 1,829 | 58.3 (55.7 to 60.8); n = 2,181 |
| Share of antibiotic-treated children who had nose-only symptoms | 72.9 (69.8 to 75.8); n = 1,048 | 64.5 (61.4 to 67.5); n = 1,244 |

Antibiotic receipt for suspected pneumonia was 62.9% in 2019 and 68.5% in 2025 (Table 2). Children with nose-only symptoms also frequently received antibiotics: 54.0% in 2019 and 54.3% in 2025 (Table 2). Their receipt was 8.8 points lower than for suspected pneumonia in 2019 (95% CI 3.0 to 14.7). By 2025, it was 14.2 points lower (95% CI 9.3 to 19.1) (Fig. 1). Because nose-only symptoms were common, these children made up 72.9% of antibiotic-treated children in 2019 and 64.5% in 2025 (Table 2).

**Fig. 1.**
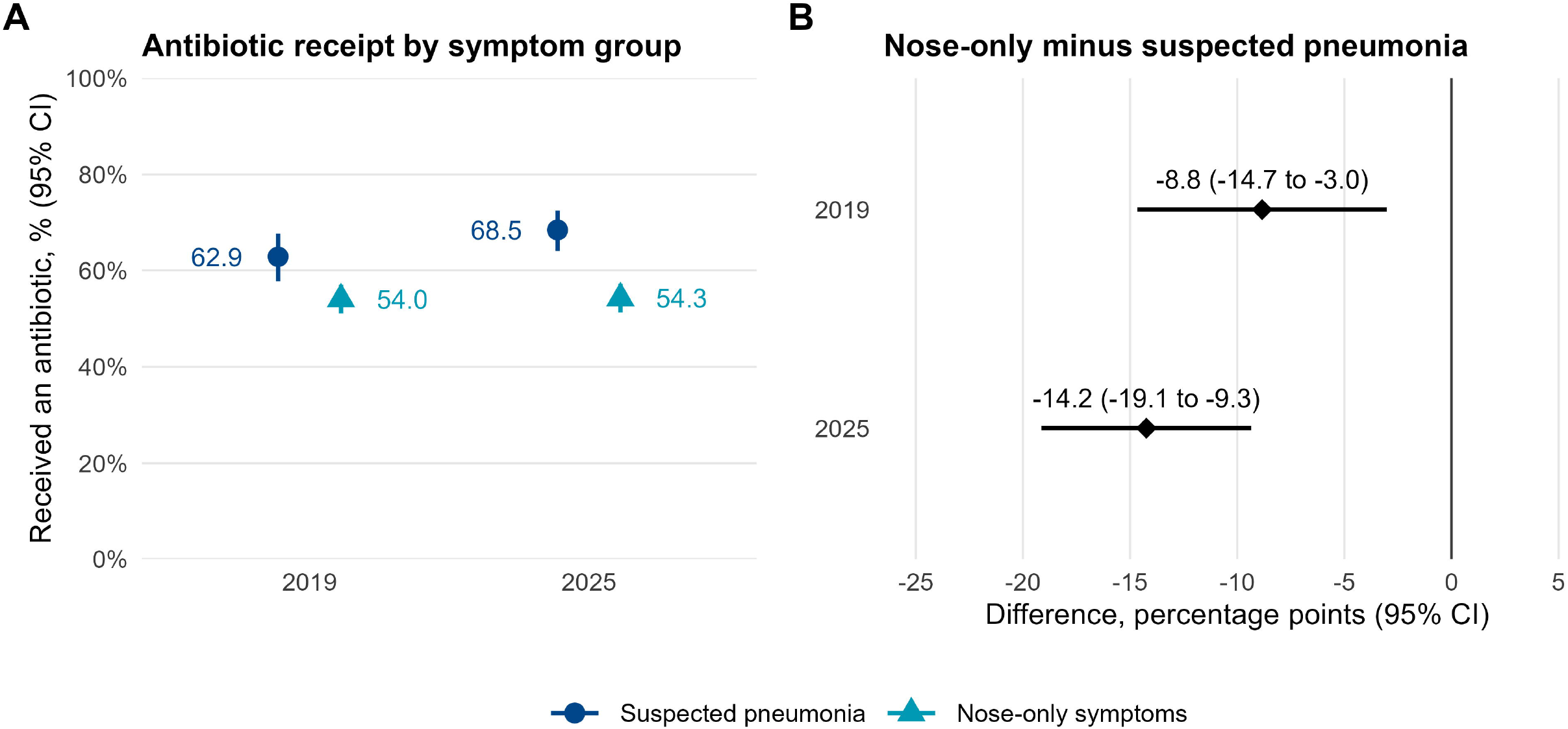
Antibiotic receipt by reported respiratory symptom group, Bangladesh MICS 2019 and 2025. (A) Survey-weighted percentage of children who received an antibiotic, with 95% CIs. Suspected pneumonia: cough with difficult breathing from the chest, or from chest and nose (n = 468 in 2019, 637 in 2025). Nose-only symptoms: cough with difficult breathing from a blocked or runny nose only (n = 1,360 and 1,526). (B) Absolute difference, nose-only minus suspected pneumonia, in percentage points with 95% CIs from survey-weighted linear models. Symptom groups are operational categories from caregiver reports; clinical need is uncertain in both, and the groups do not show whether individual courses were appropriate.

In the pooled model, the fully adjusted PR for nose-only symptoms was 0.83 (95% CI 0.78 to 0.89) (Table 3). This was a prevalence difference of -10.9 percentage points (95% CI -14.7 to -7.2). With the source of care added, the PR was 0.91 (95% CI 0.86 to 0.96), a difference of -5.7 points (95% CI -9.0 to -2.3).

**Table 3.** Antibiotic receipt by symptom location and by source of care among children with cough and difficult breathing, 2019 and 2025 pooled.

| Exposure | n | Antibiotic, % (95% CI) | Crude PR (95% CI) | Minimally adjusted PR (95% CI) | Fully adjusted PR (95% CI) | Fully adjusted prevalence difference, points (95% CI) |
|---|---|---|---|---|---|---|
| **Symptoms** |  |  |  |  |  |  |
| Suspected pneumonia (chest, or chest and nose) | 1,105 | 66.1 (62.8 to 69.3) | 1 (ref) | 1 (ref) | 1 (ref) | 0 (ref) |
| Nose only | 2,885 | 54.2 (52.1 to 56.3) | 0.82 (0.77 to 0.87) | 0.83 (0.78 to 0.88) | 0.83 (0.78 to 0.89) | -10.9 (-14.7 to -7.2) |
| Nose only, also adjusted for source of care |  |  |  |  | 0.91 (0.86 to 0.96) | -5.7 (-9.0 to -2.3) |
| **Source of care** |  |  |  |  |  |  |
| Public health care | 524 | 70.5 (65.8 to 74.8) | 1 (ref) | 1 (ref) | 1 (ref) | 0 (ref) |
| Private or NGO medical | 764 | 72.3 (68.2 to 76.0) | 1.02 (0.94 to 1.11) | 1.03 (0.95 to 1.12) | 1.04 (0.96 to 1.13) | 2.9 (-2.9 to 8.6) |
| Pharmacy, no medical facility | 1,111 | 67.8 (64.6 to 70.8) | 0.96 (0.89 to 1.04) | 0.97 (0.90 to 1.05) | 0.98 (0.91 to 1.07) | -1.1 (-6.5 to 4.4) |
| Informal provider only (no medical source) | 826 | 58.6 (54.5 to 62.6) | 0.83 (0.76 to 0.91) | 0.84 (0.76 to 0.92) | 0.86 (0.78 to 0.95) | -9.7 (-15.9 to -3.6) |
| No advice or treatment sought | 765 | 16.8 (13.9 to 20.2) | 0.24 (0.20 to 0.29) | 0.24 (0.20 to 0.29) | 0.24 (0.20 to 0.29) | -52.8 (-58.1 to -47.4) |

### Nose only

cough with difficult breathing from a blocked or runny nose only. A public, private or pharmacy source includes pharmacy care without a medical facility, so it does not mean contact with a clinician. The 1,244 treated children in 2025 include 5 with breathing difficulty from another location.

Care seeking changed between the surveys. Pharmacy care rose from 18.8% of children with cough and difficult breathing in 2019 to 36.5% in 2025 (Fig. 2A). Over the same period, informal care and no advice or treatment fell from 46.5% to 34.0% (Table S7). For suspected pneumonia, care from a public, private or pharmacy source rose from 68.2% to 77.5% (Table 2).

**Fig. 2.**
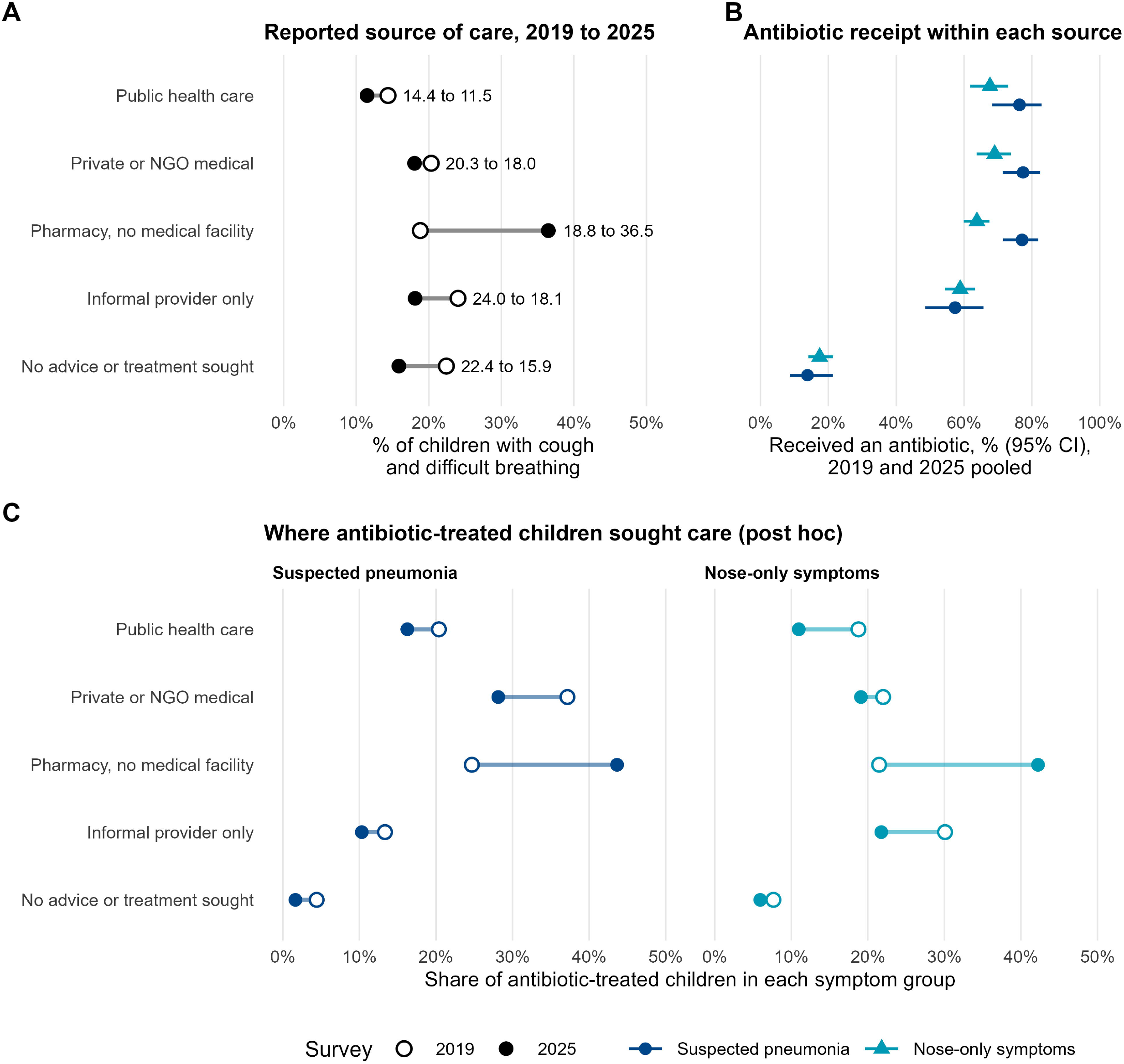
Source of care and antibiotic receipt among children with cough and difficult breathing. (A) Reported source of care in 2019 (open circles) and 2025 (filled circles), % of children with cough and difficult breathing (n = 1,829 and 2,181). One source was assigned per child in the order public health care, private or NGO medical, pharmacy, informal provider, no advice or treatment sought. Pharmacy, no medical facility: a pharmacy contact without public or private medical contact. (B) Antibiotic receipt within each source, 2019 and 2025 pooled, with 95% CIs. (C) Distribution of antibiotic-treated children across sources, 2019 and 2025, by symptom group (post hoc; numbers and CIs in Table S5). Panel C shows where observed receipt is concentrated, not inappropriate use. Source of care is where advice was sought, not who prescribed or supplied the antibiotic. All estimates are survey weighted.

Antibiotic receipt, in turn, depended strongly on whether care was sought. In the pooled sample, unadjusted receipt ranged from 72.3% with private or NGO medical care to 16.8% with no advice or treatment sought (Table 3, Fig. 2B). Against public health care, the fully adjusted PR was 1.04 (95% CI 0.96 to 1.13) for private or NGO care and 0.98 (95% CI 0.91 to 1.07) for pharmacies. It was 0.86 (95% CI 0.78 to 0.95) for informal care only and 0.24 (95% CI 0.20 to 0.29) for no advice or treatment sought.

As a result, pharmacies became a main setting for antibiotic-treated children. Among treated children with suspected pneumonia, the share with pharmacy care rose from 24.7% in 2019 to 43.7% in 2025 (Fig. 2C, Table S5, post hoc). Among treated children with nose-only symptoms, it rose from 21.5% to 42.2%. Among all children with cough and difficult breathing, those who both received an antibiotic and had pharmacy care rose from 12.6% to 24.8% (Table S5, post hoc).

Despite this shift, overall receipt changed by 2.1 percentage points (95% CI -1.6 to 5.7) (Table S8). In the decomposition, the change in where children went added 3.6 points (95% CI 2.0 to 5.3), mainly through the growth of pharmacy care. By contrast, the change in receipt within sources was -1.6 points (95% CI -5.0 to 1.9).

Across the pooled surveys, antibiotic receipt for suspected pneumonia was higher in richer children (SII 12.5 percentage points, 95% CI 1.0 to 24.0) (Fig. 3, Table S9). Care from a public, private or pharmacy source for suspected pneumonia had a larger SII (31.2, 95% CI 21.3 to 41.1), although we did not test the difference. Without care, receipt was low: among children with suspected pneumonia and no advice or treatment sought, it was 16.2% in 2019 and 10.9% in 2025 (Table S7). By contrast, receipt for nose-only symptoms showed no clear wealth gradient (SII 4.9, 95% CI -2.6 to 12.4). Neither did amoxicillin receipt for suspected pneumonia (SII 0.1, 95% CI -9.0 to 9.1; Fig. 3).

**Fig. 3.**
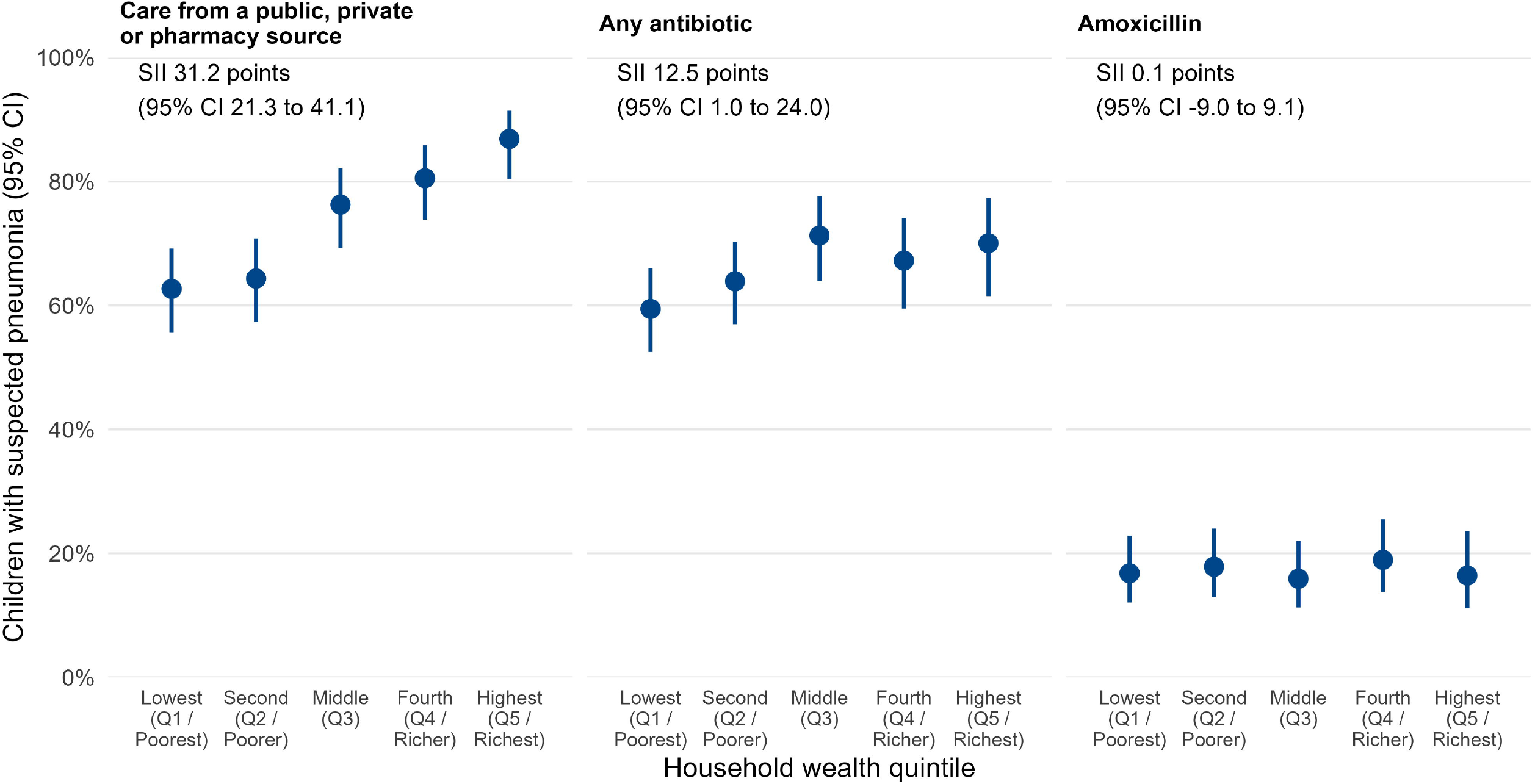
Wealth gradients in care seeking and antibiotic receipt for suspected pneumonia, 2019 and 2025 pooled (n = 1,105). Survey-weighted percentage in each wealth quintile with 95% CIs. Care from a public, private or pharmacy source includes pharmacy care without a medical facility. Amoxicillin: recorded amoxicillin, with untreated children counted as not receiving it. SII, slope index of inequality: the modelled percentage point gap between the top and bottom of the ridit-scored wealth distribution, from survey-weighted linear models adjusted for survey (Table S9).

Recorded amoxicillin receipt in suspected pneumonia was 16.6% in 2019 and 17.7% in 2025 (Table 2). In children aged 2 to 59 months it was 16.7% and 18.2% (Table S4, post hoc). Among treated children with suspected pneumonia, about one in four received amoxicillin in both years, and most received another oral antibiotic (Table S10). Among all antibiotic-treated children with cough and difficult breathing, recorded injection or IV antibiotic receipt increased from 4.6% in 2019 to 17.7% in 2025 (Table S10). This change may partly reflect recording, so we treat it as secondary.

Where advice was sought and where the antibiotic was obtained often differed. In 2019, 64.0% of treated children obtained the antibiotic from a pharmacy (Table S11). This share was 59.8% when care included public health care, 81.7% with private or NGO medical care and 63.0% with no advice or treatment sought.

The main findings were stable. Across the sensitivity analyses, the symptom PR ranged from 0.82 to 0.87, and the no-care PR changed little (Table S6). For example, adding fever to the 2019 model moved the symptom PR from 0.87 (95% CI 0.79 to 0.96) to 0.89 (95% CI 0.81 to 0.97). It changed no 2019 source PR by more than 0.01 (Table S4, post hoc). Similarly, excluding the 25 pharmacy-group children with an informal contact left the pharmacy PR at 0.98 (95% CI 0.91 to 1.06) (Table S5). For unmeasured confounding, the E-value for the symptom PR was 1.69 (1.51 for the CI bound) (Table S6).

Two results were less stable. First, the informal-care PR was 0.78 (95% CI 0.69 to 0.88) in 2019 and 0.94 (95% CI 0.81 to 1.09) in 2025 (Table S6). Second, medicines of unknown type rose from 0.7% of children in 2019 to 7.6% in 2025 (Table S6). In the unknown-type reclassification scenario, 2025 receipt rose to 76.4% for suspected pneumonia and 61.8% for nose-only symptoms (Table S6). The informal-care PR then became 0.95 (95% CI 0.88 to 1.03) (Table S6). The 2025 pharmacy share of treated children with suspected pneumonia was 41.5% in this scenario (Table S5).

## Discussion

This study asked which Bangladeshi children with cough and difficult breathing received antibiotics, where they sought care and how this differed by wealth. In 2019 and 2025, about two in three children with suspected pneumonia received an antibiotic. So did more than half of children with only nose-related breathing difficulty, who made up about two in three treated children. After adjustment, receipt for nose-only symptoms was 11 percentage points lower than for suspected pneumonia. Over the same period, care contacts shifted towards pharmacies, which accounted for more than two in five treated children by 2025. Children with no advice or treatment sought were far less likely to receive an antibiotic. Finally, receipt for suspected pneumonia and care seeking were lower in poorer children.

The first finding is that antibiotic receipt was common whatever the reported symptoms. Antibiotics give no benefit for colds and runny noses [9], yet more than half of children with nose-only symptoms received one. However, survey reports are an imperfect guide to pneumonia, so this contrast must be read with care [24,25]. In a Bangladeshi validation study against physician diagnosis, the survey questions had a sensitivity of 45.5% and a specificity of 69.5% [25]. As a result, many children in the suspected pneumonia group will not have pneumonia, and some nose-only children may have had a concurrent illness. Misclassification of symptoms or medicines may also distort the contrast, in a direction these data cannot show. The two groups are therefore operational categories with uncertain clinical need. Even so, the nose-only group was large and frequently treated.

These levels are higher than most earlier estimates, although definitions differ. Our suspected pneumonia estimates exceed the 39% (95% CI 35.4 to 42.0) found in the 2014 Bangladesh Demographic and Health Survey (DHS) [19]. However, the surveys were five or more years apart and used different questionnaires, so the gap should not be read as a trend. Al Fidah and colleagues reported 65% from the 2019 MICS [20], close to our estimate. Both estimates come from the same survey, so this agreement is not independent confirmation. Our 54% rate for nose-only symptoms is above the 33% average for non-febrile illness across 45 countries [4]. Earlier Bangladeshi studies reported 32.6% for pooled childhood illnesses [17] and 17% for fever alone [18]. A 2026 caregiver survey reported 26.3% for any respiratory illness [10].

Antibiotic type adds to this picture. Fewer than one in five children with suspected pneumonia received recorded amoxicillin, the recommended first-line drug [7]. Nepalese paediatric outpatients with respiratory infection showed a similar pattern: 64% received antibiotics, but only 9% of these received amoxicillin [36]. MICS does not name other antibiotics, but a caregiver study in Bangladesh found cefixime and azithromycin most common for respiratory illness [10]. Both are Watch antibiotics [8]. Because MICS cannot tell which children had pneumonia, low amoxicillin receipt is not a measured treatment gap. The recorded rise in injections also needs confirmation, because it could partly reflect a change in recording.

The second finding is the move of care, and of antibiotic-treated children, towards pharmacies. Adjusted receipt with pharmacy care did not clearly differ from public health care. The shift therefore changed mainly where treated children had sought care, rather than overall receipt. This describes the care pathway, not the performance of any provider, because the prescriber was not recorded. Supply without prescription is common elsewhere: in a global review, 67% of pharmacy consultations for upper respiratory symptoms ended in non-prescription antibiotic supply [37]. At the same time, receipt within sources still differed by symptom group, which may reflect some symptom-based decisions.

Where advice was sought and where the antibiotic was obtained often differed. In 2019, most treated children obtained the antibiotic from a pharmacy, including most of those whose care included public health care. Caregivers who reported no advice or treatment also mostly obtained antibiotics from pharmacies. Self-medication is one possible explanation, although prescription status was not recorded.

Qualitative work in Bangladesh describes providers and families who see antibiotics as powerful medicines for almost any illness, so caregiver demand may act alongside provider practice [14]. Village doctors are a common first contact in rural Bangladesh and often dispense antibiotics [15,38]. MICS has no code for them, so we cannot tell how caregivers recorded them. Together, these findings identify pharmacies as an important setting in which to evaluate assessment, dispensing and referral interventions.

The third finding concerns equity. Children with suspected pneumonia from poorer homes were less likely to receive an antibiotic and less likely to reach a public, private or pharmacy source. However, we did not test whether access explains the gap. Receipt for nose-only symptoms showed no clear wealth gradient, and amoxicillin receipt was low in every wealth group. In India, a national survey found antibiotic use in 18.7% of children under three with respiratory symptoms, more often with access to any source of care [39]. This resembles our pattern, although our receipt was higher. Across LMICs, recent increases in antibiotic use were largest in poor and rural groups [22]. Efforts to reduce unnecessary use should therefore be examined alongside equitable access to assessment and treatment.

This study used nationally representative surveys and reproduced the published report values. It also compared two operational symptom groups in two recent surveys, and separated where advice was sought from where antibiotics were obtained. The main findings held in several sensitivity analyses.

First, caregiver reports cannot diagnose pneumonia, so the symptom groups do not show clinical need [24,25]. Second, medicines of unknown type rose to 7.6% in 2025, so antibiotic receipt in that year may be underestimated. Third, the source of care records where advice was sought, not who prescribed or supplied the antibiotic. Fourth, the 2025 survey did not ask about fever, so confounding by illness severity may remain. Fifth, the data are cross-sectional and show associations only.

Bangladesh’s strategy for 2023 to 2028 plans to restrict over-the-counter sale of antimicrobials and to introduce stewardship and good pharmacy practice in pharmacies [13]. Such restriction has been linked to lower antibiotic use for childhood respiratory infection across countries [40]. Against this background, our findings point to three areas for evaluation rather than to proven solutions. First, clinical assessment and treatment guidance [7] could be strengthened at every source of care and monitored by receipt in each symptom group, alongside clinically assessed appropriateness. Second, pharmacies now see a large share of children with cough and difficult breathing. Dispensing guidance, prescription review and referral could be tested there. Any role for pharmacies in first-line amoxicillin would need a legal and supervised pathway, such as accredited drug shops with referral [41]. Third, poorer children were less likely to reach a public, private or pharmacy source. Without any advice or treatment, only 11% to 16% of children with suspected pneumonia received an antibiotic. Access to assessment and first-line treatment for poorer children should therefore be monitored by wealth gradients in care and treatment. Finally, future surveys could record antibiotic names, where antibiotics were obtained and how injections were given.

## Conclusions

Antibiotic receipt was common among Bangladeshi children under five with reported cough and difficult breathing in both surveys, including those with nose-only symptoms. Reported care contacts shifted towards pharmacies, while receipt and care seeking differed across wealth groups. These findings support monitoring antibiotic receipt alongside symptoms and care contacts. They also support evaluating stewardship across medical and pharmacy settings while protecting access to assessment and appropriate treatment. The surveys cannot determine the appropriateness of individual antibiotic courses.

## Data Availability

The MICS datasets analysed in this study are available from the UNICEF MICS website (https://mics.unicef.org) after registration and approval of a data request.

https://mics.unicef.org/surveys

## Abbreviations

CI: confidence interval
DHS: Demographic and Health Survey
IV: intravenous
LMICs: low-income and middle-income countries
MICS: Multiple Indicator Cluster Survey
NGO: non-governmental organisation
PR: prevalence ratio
SII: slope index of inequality
STROBE: Strengthening the Reporting of Observational Studies in Epidemiology
UNICEF: United Nations Children’s Fund
WHO: World Health Organization.

## Declarations

### Ethics approval and consent to participate

This secondary analysis used anonymised data obtained from the UNICEF MICS programme. The 2025 survey protocol was approved by the Ethical Review Committee of icddr,b (International Centre for Diarrhoeal Disease Research, Bangladesh) in November 2024 (reference PR-24113) [30]. The 2019 protocol was reviewed and approved by a Government of Bangladesh technical committee under the Bangladesh Bureau of Statistics [29]. The 2019 and 2025 reports state that respondents gave informed consent [29,30].

### Availability of data and materials

The MICS datasets analysed in this study are available from the UNICEF MICS website (https://mics.unicef.org) after registration and approval of a data request. Aggregate estimates are in this article and Additional file 1.

## Acknowledgements

We thank the Bangladesh Bureau of Statistics and UNICEF for making the MICS data available.

