## Supplementary file for "Antibiotic receipt by respiratory symptom group and source of care among children under five in Bangladesh: repeated cross-sectional analyses of national surveys, 2019 and 2025"

Additional file 1. Supplementary tables

All estimates are survey weighted unless stated otherwise and were produced in Stata version 17. ARI, acute respiratory infection; CI, confidence interval; df, degrees of freedom; IV, intravenous; NGO, non-governmental organisation; PR, prevalence ratio; SII, slope index of inequality; WHO, World Health Organization.

**Table S1. Harmonisation of questionnaire items across surveys (A), candidate indicators for monitoring antibiotic use in sick children (B) and medicine codes recorded in 2019 and 2025 (C).**

Panel A: item identifiers and question wording from each round’s under-five questionnaire, and the children asked each item. Panel B: proposed indicators; all need validation, and none measures the appropriateness of an individual course. Amoxicillin receipt is not the same as the Access share of antibiotics. Panel C: unweighted numbers of children with a positive child weight given any medicine for the illness, and the number with each code. Universes differ: fever or cough in 2019, difficult breathing in 2025. Several codes could be recorded per child. In 2019 paracetamol was recorded far more often than aspirin; in 2025 the reverse held, which may be a coding or recording discrepancy. The antibiotic codes (L to O) reproduce the published 2025 estimate. This supports consistency with the report but does not independently validate medicine classification or exclude recording errors in antibiotic categories. Non-antibiotic medicines were not analysed.

| Item | 2006 | 2012-13 | 2019 | 2025 |
| --- | --- | --- | --- | --- |
| Cough in the last 2 weeks | CA5 | CA7 | CA16 | AR1 |
| Difficult breathing | CA6, asked if cough | CA8, asked if cough | CA17, asked of all children | AR2, asked of all children |
| Breathing problem in chest, nose or both | CA7 | CA9 | CA18 | AR3 |
| Fever in the last 2 weeks | Not used | Not used | CA14 | Not asked |
| Sought advice or treatment; places (multiple answers) | CA8; CA9A to CA9X | CA10; CA11A to CA11X | CA20; CA21A to CA21X | AR4; AR5A to AR5Z |
| Medicine given: universe | CA10; chest, chest and nose, or don’t know | CA12; chest, chest and nose, or don’t know | CA22; fever or cough (all children with cough and difficult breathing) | AR9; difficult breathing |
| Medicine question wording | CA10, was (name) given medicine to treat this illness? | CA12, was (name) given any medicine to treat this illness? | CA22, at any time during the illness, was (name) given any medicine for the illness? | AR9, at any time during the illness, did (name) take any medicine for the illness? |
| Paracetamol; aspirin | CA11P; CA11Q | CA13P; CA13Q | CA23R; CA23S | AR10R; AR10S |
| Antibiotic items | CA11A any antibiotic | CA13A pill or syrup; CA13B injection | CA23L amoxicillin; CA23M cotrimoxazole; CA23N other pill or syrup; CA23O other injection or IV | AR10L; AR10M; AR10N; AR10O (same labels as 2019) |
| Only brand name recorded | No code | No code | CA23W | AR10W |
| Medicine type not known | CA11Z | CA13Z | CA23Z | AR10Z |
| Where the antibiotic was obtained | Not asked | Not asked | CA25A to CA25X | Not asked |
| Diarrhoea in the last 2 weeks | CA1 | CA1 | CA1 | DA1 |
| Gate question: anything else given for diarrhoea | Not asked | CA5 | CA12 | DA10 |
| Diarrhoea antibiotic (pill or syrup; injection) | Not asked | CA6A; CA6L | CA13A; CA13L | DA11A; DA11L |
| Antimotility drug | Not asked | CA6B | CA13B | DA11B |

| Dimension | Candidate indicator | Denominator | Rounds | Main limitation |
| --- | --- | --- | --- | --- |
| Symptom pattern | Antibiotic receipt for chest-related and for nose-only symptoms | Children with cough and difficult breathing in each symptom group | 2019, 2025 | Caregiver report; not a clinical diagnosis |
| Care seeking | Reported source of care, five groups | Children with cough and difficult breathing | 2019, 2025 | Advice location, not the prescriber |
| Concentration of receipt | Distribution of antibiotic-treated children across sources | Antibiotic-treated children in each symptom group | 2019, 2025 | Depends on source priority rule |
| Medicine acquisition | Where the antibiotic was obtained | Antibiotic-treated children | 2019 | Multiple answers; asked in one round |
| Medicine selection | Recorded amoxicillin; recorded injection or IV antibiotic | Children with suspected pneumonia; antibiotic-treated children | 2019, 2025 | Other antibiotics not named |
| Equity | Slope index of inequality in care and antibiotic receipt | Children in each symptom group | 2019, 2025 | Linear summary of quintiles |
| Measurement quality | Medicines of unknown type; brand name only | Children with cough and difficult breathing | 2019, 2025 | Reclassification is not an exhaustive upper bound |

| Medicine code | 2019 (CA23), n | 2025 (AR10), n |
| --- | --- | --- |
| Children given any medicine (denominator) | 5,019 | 2,059 |
| Amoxicillin (L) | 470 | 347 |
| Cotrimoxazole (M) | 87 | 71 |
| Other antibiotic pill or syrup (N) | 2,067 | 839 |
| Other antibiotic injection or IV (O) | 90 | 230 |
| Paracetamol (R) | 2,936 | 35 |
| Aspirin (S) | 29 | 670 |
| Ibuprofen (T) | 50 | 38 |
| Only brand name recorded (W) | 53 | 23 |
| Medicine type not known (Z) | 36 | 190 |

**Table S2. Antibiotic and antimotility estimates compared with the published MICS reports (%).**

Report tables: 2006 CH.7; 2012-13 CH.6 and CH.5; 2019 TC.4.1 and TC.4.4; 2025 TC.5.1 and TC.4.4. The 2025 report defines ARI symptoms as difficult breathing from the chest, without requiring cough. Report diarrhoea values are given by route; this analysis combines them.

| Indicator | Survey | Published report | This analysis | Unweighted n | Weighted n |
| --- | --- | --- | --- | --- | --- |
| Antibiotic, ARI symptoms (report definition) | 2006 | 21.5 | 21.5 | 1,598 | 1,683 |
| Antibiotic, ARI symptoms (report definition) | 2012-13 | 74.3 | 74.3 | 711 | 670 |
| Antibiotic, ARI symptoms (report definition) | 2019 | 62.9 | 62.9 | 468 | 470 |
| Antibiotic, ARI symptoms (report definition) | 2025 | 68.6 | 68.6 | 671 | 671 |
| Antibiotic, diarrhoea | 2006 | Not asked | Not asked |  |  |
| Antibiotic, diarrhoea | 2012-13 | 18.8 pill and 1.4 injection | 20.1 | 829 | 825 |
| Antibiotic, diarrhoea | 2019 | 5.2 pill and 0.5 injection | 5.6 | 1,588 | 1,596 |
| Antibiotic, diarrhoea | 2025 | 13.3 | 13.3 | 1,634 | 1,651 |
| Antimotility, diarrhoea | 2006 | Not asked | Not asked |  |  |
| Antimotility, diarrhoea | 2012-13 | 2.4 | 2.4 | 829 |  |
| Antimotility, diarrhoea | 2019 | 7.6 | 7.6 | 1,588 |  |
| Antimotility, diarrhoea | 2025 | 19.0 | 19.0 | 1,634 |  |

**Table S3. Sample sizes by survey, exclusions from the model sample and mixed care contacts. Characteristics of the model sample are in Table 1.**

Counts are unweighted numbers of children. Suspected pneumonia, nose only and other location are subgroups of cough with difficult breathing. Antibiotic use for diarrhoea was not asked in 2006. Among the 4,010 children with cough and difficult breathing in 2019 and 2025, 19 had breathing difficulty from another or unknown location and 1 had missing diarrhoea data. No other model variable had missing values, so the model sample excludes 20 children. Pharmacy, no medical facility: a pharmacy contact without public or private medical contact; 17 children in 2019 and 8 in 2025 in this group also reported an informal provider. Higher-priority groups can include lower-level contacts. Among care-seekers, 49 children in 2019 and 28 in 2025 in the public group had another contact. In the private group, 39 and 27 had a pharmacy or informal contact. Children are counted when the under-five questionnaire was completed (positive child weight). In 2019 one further child with diarrhoea had a partly completed questionnaire and a weight of zero; analyses that count every valid diarrhoea answer give 1,589 instead of 1,588.

| Group | 2006 | 2012-13 | 2019 | 2025 |
| --- | --- | --- | --- | --- |
| Children under five with a completed questionnaire | 31,566 | 20,903 | 23,099 | 23,357 |
| Cough with difficult breathing | 3,638 | 3,260 | 1,829 | 2,181 |
| Suspected pneumonia (chest, or chest and nose) | 1,598 | 711 | 468 | 637 |
| Nose only | 1,970 | 2,501 | 1,360 | 1,526 |
| Other or unknown location of breathing difficulty | 70 | 48 | 1 | 18 |
| Diarrhoea | 2,250 | 829 | 1,588 | 1,634 |
| Diarrhoea with antibiotic use measured | Not asked | 829 | 1,588 | 1,634 |
| Model sample (cough with difficult breathing, complete data) |  |  | 1,827 | 2,163 |

**Table S4. Post hoc checks: fever in 2019, age range of WHO pneumonia guidance, fitted and counterfactual probabilities, logistic standardisation and survey design.**

Fever was asked in 2019 only, so the fever checks are restricted to 2019. Design: Taylor linearisation, cluster identifiers unique within each survey, strata survey specific and single-cluster strata centred. Age check: children aged 2 to 59 months, the range of WHO guidance; 17 children in 2019 and 19 in 2025 with suspected pneumonia were under 2 months. Counterfactual predictions: every child in the model sample set to one exposure level, as used for the standardised prevalence differences in Table 3. Logistic rows: marginal differences in percentage points from survey-weighted logistic models with the same covariates, for comparison with the Poisson-based differences in Table 3. Neither check establishes whether an individual course was appropriate. These analyses were not planned in advance.

| Check | Assumption tested | Estimate (95% CI) |
| --- | --- | --- |
| 2019 symptom PR, fully adjusted (as in Table 3, 2019 only) | Reference | 0.87 (0.79 to 0.96); n = 1,827 |
| 2019 symptom PR, fever added | Confounding by fever (not asked in 2025) | 0.89 (0.81 to 0.97) |
| Children in the 2019 model sample with fever, % |  | 63.8 |
| Amoxicillin, suspected pneumonia, all ages, 2019 | Reference | 16.6 (13.0 to 20.9); n = 468 |
| Amoxicillin, suspected pneumonia, 2 to 59 months, 2019 | Age range of WHO pneumonia guidance | 16.7 (13.0 to 21.2); n = 451 |
| Amoxicillin, suspected pneumonia, all ages, 2025 | Reference | 17.7 (14.7 to 21.1); n = 637 |
| Amoxicillin, suspected pneumonia, 2 to 59 months, 2025 | Age range of WHO pneumonia guidance | 18.2 (15.1 to 21.7); n = 618 |
| Fitted probabilities above 1, source of care model | Prevalence differences from a Poisson model | 5 of 3,990 (maximum 1.05) |
| Fitted probabilities above 1, symptom model | Prevalence differences from a Poisson model | 0 of 3,990 (maximum 0.95) |
| Counterfactual predictions above 1, all children set to public health care | Standardised differences, source model | 4 of 3,990 (maximum 1.01) |
| Counterfactual predictions above 1, all children set to private or NGO medical | Standardised differences, source model | 11 of 3,990 (maximum 1.05) |
| Counterfactual predictions above 1, all children set to pharmacy, no medical facility | Standardised differences, source model | 0 of 3,990 (maximum 0.99) |
| Counterfactual predictions above 1, all children set to informal provider | Standardised differences, source model | 0 of 3,990 (maximum 0.87) |
| Counterfactual predictions above 1, all children set to no advice or treatment sought | Standardised differences, source model | 0 of 3,990 (maximum 0.24) |
| Counterfactual predictions above 1, all children set to suspected pneumonia | Standardised differences, symptom model | 0 of 3,990 (maximum 0.98) |
| Counterfactual predictions above 1, all children set to nose only | Standardised differences, symptom model | 0 of 3,990 (maximum 0.81) |
| Logistic marginal difference, nose only vs suspected pneumonia, points | Model for standardised differences | -11.0 (-14.8 to -7.2) |
| Logistic marginal difference, private or NGO medical vs public health care, points | Model for standardised differences | 2.8 (-3.0 to 8.6) |
| Logistic marginal difference, pharmacy, no medical facility vs public health care, points | Model for standardised differences | -1.3 (-6.8 to 4.3) |
| Logistic marginal difference, informal provider vs public health care, points | Model for standardised differences | -9.6 (-15.8 to -3.4) |
| Logistic marginal difference, no advice or treatment sought vs public health care, points | Model for standardised differences | -53.1 (-58.5 to -47.7) |
| 2019 source PR, private or NGO medical vs public health care, fully adjusted | Reference | 1.00 (0.90 to 1.11); n = 1,827 |
| 2019 source PR, private or NGO medical vs public health care, fever added | Confounding by fever (not asked in 2025) | 0.99 (0.90 to 1.10) |
| 2019 source PR, pharmacy, no medical facility vs public health care, fully adjusted | Reference | 0.91 (0.81 to 1.02); n = 1,827 |
| 2019 source PR, pharmacy, no medical facility vs public health care, fever added | Confounding by fever (not asked in 2025) | 0.91 (0.81 to 1.02) |
| 2019 source PR, informal provider vs public health care, fully adjusted | Reference | 0.78 (0.69 to 0.88); n = 1,827 |
| 2019 source PR, informal provider vs public health care, fever added | Confounding by fever (not asked in 2025) | 0.78 (0.69 to 0.88) |
| 2019 source PR, no advice or treatment sought vs public health care, fully adjusted | Reference | 0.22 (0.17 to 0.29); n = 1,827 |
| 2019 source PR, no advice or treatment sought vs public health care, fever added | Confounding by fever (not asked in 2025) | 0.22 (0.17 to 0.29) |
| Survey design of the pooled model sample | Variance estimation | 251 strata; 6,291 clusters; 6,040 design df |

**Table S5. Post hoc checks of the pharmacy findings: excluding mixed pharmacy and informal contacts (A), pharmacy share of treated children (B) and joint receipt by source (C).**

Pharmacy-group children with an informal provider were 6.5% (4.1 to 10.1) of the pharmacy group in 2019 (n = 324) and 1.0% (0.5 to 2.3) in 2025 (n = 792). Panel A: fully adjusted source model as in Table 3. Panel B: % of antibiotic-treated children in each symptom group who had pharmacy care without a medical facility. In the last column, children given a medicine of unknown type also count as treated. Panel C: the denominator is all children with cough and difficult breathing in each survey. Each cell is the % who received an antibiotic and had that source, so the cells sum to overall receipt. Panel C shows where observed receipt is concentrated; it is not a preventable fraction and does not measure inappropriate use. These analyses were not planned in advance.

| Source of care (reference public health care) | Primary analysis, PR (95% CI); n = 3,990 | Excluding pharmacy-group children with an informal contact, PR (95% CI); n = 3,965 |
| --- | --- | --- |
| Private or NGO medical | 1.04 (0.96 to 1.13) | 1.04 (0.96 to 1.13) |
| Pharmacy, no medical facility | 0.98 (0.91 to 1.07) | 0.98 (0.91 to 1.06) |
| Informal provider | 0.86 (0.78 to 0.95) | 0.86 (0.78 to 0.95) |
| No advice or treatment sought | 0.24 (0.20 to 0.29) | 0.24 (0.20 to 0.29) |

| Symptom group | Survey | Primary analysis | Excluding pharmacy-group children with an informal contact | Unknown-type medicines counted as antibiotics |
| --- | --- | --- | --- | --- |
| Suspected pneumonia | 2019 | 24.7 (19.4 to 30.9); n = 303 | 24.1 (18.8 to 30.3); n = 301 | 24.5 (19.2 to 30.6); n = 308 |
| Suspected pneumonia | 2025 | 43.7 (38.5 to 48.9); n = 429 | 42.9 (37.8 to 48.2); n = 424 | 41.5 (36.7 to 46.5); n = 480 |
| Nose only | 2019 | 21.5 (18.2 to 25.2); n = 745 | 20.4 (17.2 to 24.1); n = 737 | 21.4 (18.1 to 25.1); n = 755 |
| Nose only | 2025 | 42.2 (38.3 to 46.3); n = 810 | 42.0 (38.1 to 46.1); n = 807 | 39.9 (36.3 to 43.6); n = 936 |

| Source of care | 2019 | 2025 |
| --- | --- | --- |
| Public health care | 10.8 (9.3 to 12.5); n = 1,829 | 7.5 (6.3 to 8.9); n = 2,181 |
| Private or NGO medical | 14.7 (12.9 to 16.6); n = 1,829 | 13.0 (11.5 to 14.7); n = 2,181 |
| Pharmacy, no medical facility | 12.6 (10.9 to 14.4); n = 1,829 | 24.8 (22.7 to 26.9); n = 2,181 |
| Informal provider | 14.4 (12.7 to 16.2); n = 1,829 | 10.3 (8.8 to 12.0); n = 2,181 |
| No advice or treatment sought | 3.8 (2.9 to 5.0); n = 1,829 | 2.6 (1.9 to 3.5); n = 2,181 |

**Table S6. Sensitivity analyses for the two exposures: fully adjusted prevalence ratios (95% CI), each analysis changing one assumption.**

Reference groups: suspected pneumonia; public health care. Chest-only reference group: nose only compared with chest-only symptoms, in a model with chest and nose as a separate third level. Logistic rows are odds ratios. Interview season: winter (December to February), summer (March to May), monsoon (June to September), post-monsoon (October and November). Unknown-type reclassification scenario: only the ‘don’t know’ medicine code was counted, not other or brand-name codes, so it is not an exhaustive upper bound. The interaction p values test whether the exposure association differs between 2019 and 2025 (symptom: 1 df; source: 4 df, joint). E-value: the minimum PR an unmeasured confounder would need with both exposure and outcome to explain away the estimate. E-values hold only under their own assumptions and do not address misclassification or selection. The second value is for the CI bound nearest 1 (1.00 when the CI includes 1). Second panel: medicines of unknown type and antibiotic receipt under the primary definition and the unknown-type reclassification scenario, % (95% CI).

| Analysis | Assumption tested | Nose only vs suspected pneumonia | Pharmacy, no medical facility vs public health care | Informal provider vs public health care | No advice or treatment sought vs public health care |
| --- | --- | --- | --- | --- | --- |
| Primary analysis (fully adjusted, Table 3) |  | 0.83 (0.78 to 0.89) | 0.98 (0.91 to 1.07) | 0.86 (0.78 to 0.95) | 0.24 (0.20 to 0.29) |
| Brand-name-only medicines counted as antibiotics | Outcome definition | 0.83 (0.78 to 0.88) | 0.99 (0.92 to 1.07) | 0.87 (0.79 to 0.95) | 0.24 (0.20 to 0.29) |
| Medicines of unknown type counted as antibiotics (unknown-type reclassification scenario) | Outcome definition | 0.84 (0.79 to 0.88) | 0.96 (0.90 to 1.04) | 0.95 (0.88 to 1.03) | 0.26 (0.22 to 0.31) |
| Logistic regression, odds ratio | Effect measure | 0.63 (0.53 to 0.74) | 0.94 (0.72 to 1.23) | 0.65 (0.48 to 0.86) | 0.08 (0.06 to 0.11) |
| Interview season added | Confounder set | 0.83 (0.78 to 0.89) | 0.99 (0.91 to 1.07) | 0.86 (0.78 to 0.95) | 0.24 (0.20 to 0.29) |
| Children without diarrhoea | Co-illness | 0.84 (0.78 to 0.89) |  |  |  |
| 2019 only | Pooling of surveys | 0.87 (0.79 to 0.96) | 0.91 (0.81 to 1.02) | 0.78 (0.69 to 0.88) | 0.22 (0.17 to 0.29) |
| 2025 only | Pooling of surveys | 0.82 (0.76 to 0.89) | 1.07 (0.95 to 1.19) | 0.94 (0.81 to 1.09) | 0.27 (0.20 to 0.36) |
| Interaction with survey, p (symptom: 1 df; source: 4 df, joint) | Pooling of surveys | 0.185 | 0.244 |  |  |
| Chest-only reference group | Exposure definition | 0.87 (0.80 to 0.94) |  |  |  |
| Four-level source (informal provider and no advice or treatment sought combined) | Exposure definition |  |  | Combined: 0.55 (0.50 to 0.61) |  |
| E-value for the fully adjusted PR | No unmeasured confounding | 1.69 (CI bound 1.51) | 1.14 (CI bound 1.00) | 1.60 (CI bound 1.30) | 7.85 (CI bound 6.34) |

| Measure | 2019 | 2025 |
| --- | --- | --- |
| Medicine of unknown type, all children with cough and difficult breathing | 0.7 (0.4 to 1.2) | 7.6 (6.5 to 9.0) |
| Suspected pneumonia: antibiotic (primary definition) | 62.9 (57.7 to 67.8) | 68.5 (64.2 to 72.5) |
| Suspected pneumonia: antibiotic or medicine of unknown type (unknown-type reclassification scenario) | 63.9 (58.8 to 68.7) | 76.4 (72.3 to 80.0) |
| Nose only: antibiotic (primary definition) | 54.0 (51.1 to 57.0) | 54.3 (51.3 to 57.2) |
| Nose only: antibiotic or medicine of unknown type (unknown-type reclassification scenario) | 54.6 (51.6 to 57.6) | 61.8 (58.8 to 64.7) |

**Table S7. Source of care by respiratory symptom group and survey (A) and antibiotic receipt by source (B), % (95% CI).**

Source assigned in the order public health care, private or NGO medical, pharmacy, informal provider, no advice or treatment sought. Informal providers: traditional practitioners, relatives, shops and other places. Pharmacy, no medical facility: a pharmacy contact without public or private medical contact; 17 children in 2019 and 8 in 2025 in this group also reported an informal provider. Higher-priority groups can include lower-level contacts.

| Group | Source | 2019 | 2025 |
| --- | --- | --- | --- |
| Cough and difficult breathing | Public health care | 14.4 (12.7 to 16.3) | 11.5 (10.0 to 13.1) |
| Cough and difficult breathing | Private or NGO medical | 20.3 (18.3 to 22.6) | 18.0 (16.2 to 20.0) |
| Cough and difficult breathing | Pharmacy, no medical facility | 18.8 (16.9 to 20.9) | 36.5 (34.2 to 38.8) |
| Cough and difficult breathing | Informal provider | 24.0 (21.9 to 26.4) | 18.1 (16.2 to 20.1) |
| Cough and difficult breathing | No advice or treatment sought | 22.4 (20.3 to 24.7) | 15.9 (14.1 to 17.8) |
| Suspected pneumonia | Public health care | 16.1 (12.7 to 20.2) | 15.1 (12.2 to 18.6) |
| Suspected pneumonia | Private or NGO medical | 30.3 (25.8 to 35.3) | 24.8 (21.2 to 28.8) |
| Suspected pneumonia | Pharmacy, no medical facility | 21.8 (17.6 to 26.6) | 37.6 (33.4 to 42.0) |
| Suspected pneumonia | Informal provider | 14.7 (11.6 to 18.5) | 12.2 (9.6 to 15.4) |
| Suspected pneumonia | No advice or treatment sought | 17.1 (13.5 to 21.3) | 10.3 (7.8 to 13.3) |
| Nose only | Public health care | 13.8 (11.9 to 16.0) | 9.9 (8.3 to 11.8) |
| Nose only | Private or NGO medical | 17.1 (14.9 to 19.6) | 15.1 (13.1 to 17.4) |
| Nose only | Pharmacy, no medical facility | 17.9 (15.7 to 20.3) | 36.2 (33.5 to 39.0) |
| Nose only | Informal provider | 27.0 (24.4 to 29.8) | 20.6 (18.3 to 23.2) |
| Nose only | No advice or treatment sought | 24.1 (21.6 to 26.9) | 18.1 (15.8 to 20.6) |

| Group | Source | 2019 | 2025 |
| --- | --- | --- | --- |
| Cough and difficult breathing | Public health care | 75.0 (68.6 to 80.5); n = 264 | 65.5 (58.6 to 71.9); n = 263 |
| Cough and difficult breathing | Private or NGO medical | 72.1 (66.4 to 77.2); n = 379 | 72.1 (66.1 to 77.5); n = 388 |
| Cough and difficult breathing | Pharmacy, no medical facility | 66.7 (61.0 to 71.9); n = 324 | 67.9 (64.0 to 71.5); n = 792 |
| Cough and difficult breathing | Informal provider | 59.7 (54.2 to 65.0); n = 460 | 57.0 (50.8 to 63.0); n = 369 |
| Cough and difficult breathing | No advice or treatment sought | 17.0 (13.1 to 21.7); n = 402 | 16.6 (12.4 to 21.7); n = 369 |
| Suspected pneumonia | Public health care | 79.7 (67.6 to 88.0); n = 80 | 73.8 (62.4 to 82.7); n = 98 |
| Suspected pneumonia | Private or NGO medical | 77.1 (68.0 to 84.2); n = 143 | 77.7 (69.2 to 84.4); n = 163 |
| Suspected pneumonia | Pharmacy, no medical facility | 71.3 (59.3 to 81.0); n = 94 | 79.6 (73.3 to 84.6); n = 234 |
| Suspected pneumonia | Informal provider | 57.0 (44.4 to 68.7); n = 73 | 57.7 (45.4 to 69.1); n = 76 |
| Suspected pneumonia | No advice or treatment sought | 16.2 (8.9 to 27.7); n = 78 | 10.9 (5.1 to 21.8); n = 66 |
| Nose only | Public health care | 73.3 (65.5 to 79.9); n = 184 | 59.9 (51.0 to 68.2); n = 162 |
| Nose only | Private or NGO medical | 69.5 (62.2 to 75.9); n = 235 | 68.5 (60.6 to 75.5); n = 223 |
| Nose only | Pharmacy, no medical facility | 64.9 (58.0 to 71.2); n = 230 | 63.3 (58.6 to 67.8); n = 553 |
| Nose only | Informal provider | 60.2 (54.2 to 65.9); n = 387 | 57.2 (50.3 to 63.8); n = 290 |
| Nose only | No advice or treatment sought | 17.2 (12.8 to 22.6); n = 324 | 17.8 (13.1 to 23.9); n = 298 |

**Table S8. Kitagawa decomposition of the change in antibiotic receipt from 2019 to 2025 among children with cough and difficult breathing, across five sources of care.**

Composition: change due to where children went. Rate: change due to antibiotic receipt within each source. Each component weights the change in one factor by the average of the other across the two surveys. Shares and rates were estimated jointly for both surveys, and CIs come from the delta method with their full survey covariance. Descriptive, not causal.

| Component | Percentage points (95% CI) |
| --- | --- |
| Total change in antibiotic use | 2.1 (-1.6 to 5.7) |
| Change in where children went (composition) | 3.6 (2.0 to 5.3) |
| Change in antibiotic use within each source (rate) | -1.6 (-5.0 to 1.9) |
| Composition: public health care | -2.0 (-3.7 to -0.4) |
| Composition: private or NGO medical | -1.7 (-3.7 to 0.4) |
| Composition: pharmacy, no medical facility | 11.9 (9.8 to 14.0) |
| Composition: informal provider | -3.5 (-5.2 to -1.7) |
| Composition: no advice or treatment sought | -1.1 (-1.6 to -0.6) |
| Rate: public health care | -1.2 (-2.4 to -0.1) |
| Rate: private or NGO medical | 0.0 (-1.5 to 1.5) |
| Rate: pharmacy, no medical facility | 0.3 (-1.5 to 2.2) |
| Rate: informal provider | -0.6 (-2.3 to 1.1) |
| Rate: no advice or treatment sought | -0.1 (-1.3 to 1.1) |

**Table S9. Antibiotic receipt and care from a public, private or pharmacy source by wealth quintile in each respiratory symptom group, with slope index of inequality (SII), 2019 and 2025.**

SII: difference in percentage points between the richest and poorest ends of the ridit-scored wealth scale, from survey-weighted linear models. Ridit scores are midpoints of weighted cumulative quintile shares within each group and survey; pooled models adjust for survey and assume a common slope in both surveys. Differences between SIIs were not tested. For care, quintile percentages are shown for the pooled surveys only.

| Indicator | Survey | Lowest (Quintile 1 / Poorest) | Second (Quintile 2 / Poorer) | Middle (Quintile 3) | Fourth (Quintile 4 / Richer) | Highest (Quintile 5 / Richest) | SII, points (95% CI) | p |
| --- | --- | --- | --- | --- | --- | --- | --- | --- |
| Suspected pneumonia: antibiotic | 2019 | 55.6 | 60.8 | 70.9 | 67.2 | 62.6 | 11.6 (-7.4 to 30.6) | 0.231 |
| Suspected pneumonia: antibiotic | 2025 | 62.7 | 66.3 | 71.6 | 67.3 | 76.2 | 13.1 (-1.1 to 27.4) | 0.070 |
| Suspected pneumonia: antibiotic | 2019 and 2025 | 59.4 | 63.9 | 71.3 | 67.2 | 70.0 | 12.5 (1.0 to 24.0) | 0.033 |
| Nose only: antibiotic | 2019 | 51.1 | 54.8 | 54.9 | 56.1 | 53.9 | 3.8 (-6.9 to 14.4) | 0.489 |
| Nose only: antibiotic | 2025 | 49.5 | 57.9 | 55.2 | 49.8 | 60.6 | 6.0 (-4.7 to 16.7) | 0.270 |
| Nose only: antibiotic | 2019 and 2025 | 50.3 | 56.3 | 55.0 | 52.8 | 56.8 | 4.9 (-2.6 to 12.4) | 0.203 |
| Suspected pneumonia: care from a public, private or pharmacy source | 2019 |  |  |  |  |  | 45.6 (30.7 to 60.5) | <0.001 |
| Suspected pneumonia: care from a public, private or pharmacy source | 2025 |  |  |  |  |  | 20.7 (7.6 to 33.8) | 0.002 |
| Suspected pneumonia: care from a public, private or pharmacy source | 2019 and 2025 | 62.7 | 64.4 | 76.3 | 80.5 | 86.9 | 31.2 (21.3 to 41.1) | <0.001 |

**Table S10. Type of antibiotic by respiratory symptom group, 2019 and 2025: overlapping types (A) and mutually exclusive patterns (B), % (95% CI).**

Panel A: categories are not exclusive, because a child could receive more than one antibiotic. Injection: recorded injection or IV antibiotic. Panel B: antibiotic-treated children, patterns assigned in the priority order shown, so each column sums to 100% within a group.

| Group | Measure | 2019 | 2025 |
| --- | --- | --- | --- |
| Suspected pneumonia | Amoxicillin, among antibiotic-treated | 26.4 (20.9 to 32.6); n = 303 | 25.8 (21.5 to 30.6); n = 429 |
| Suspected pneumonia | Cotrimoxazole, among antibiotic-treated | 3.8 (2.0 to 7.0); n = 303 | 7.6 (5.3 to 11.0); n = 429 |
| Suspected pneumonia | Other oral antibiotic, among antibiotic-treated | 72.2 (65.8 to 77.8); n = 303 | 65.2 (59.9 to 70.2); n = 429 |
| Suspected pneumonia | Injection or IV, among antibiotic-treated | 9.6 (6.5 to 14.1); n = 303 | 19.2 (15.2 to 24.0); n = 429 |
| Suspected pneumonia | Two or more antibiotic types, among antibiotic-treated | 11.3 (7.8 to 16.0); n = 303 | 15.9 (12.3 to 20.4); n = 429 |
| Suspected pneumonia | Amoxicillin, among all children in the group | 16.6 (13.0 to 20.9); n = 468 | 17.7 (14.7 to 21.1); n = 637 |
| Suspected pneumonia | Brand name only recorded, among children without an antibiotic | 3.6 (1.4 to 9.0); n = 165 | 2.5 (0.7 to 8.4); n = 208 |
| Nose only | Amoxicillin, among antibiotic-treated | 16.5 (13.7 to 19.8); n = 745 | 25.1 (21.6 to 28.9); n = 810 |
| Nose only | Cotrimoxazole, among antibiotic-treated | 3.6 (2.4 to 5.4); n = 745 | 4.1 (2.6 to 6.4); n = 810 |
| Nose only | Other oral antibiotic, among antibiotic-treated | 81.8 (78.5 to 84.8); n = 745 | 64.0 (59.9 to 68.0); n = 810 |
| Nose only | Injection or IV, among antibiotic-treated | 2.8 (1.8 to 4.3); n = 745 | 16.8 (13.3 to 20.9); n = 810 |
| Nose only | Two or more antibiotic types, among antibiotic-treated | 4.6 (3.0 to 6.9); n = 745 | 8.7 (6.6 to 11.4); n = 810 |
| Nose only | Amoxicillin, among all children in the group | 8.9 (7.3 to 10.8); n = 1,360 | 13.6 (11.6 to 15.9); n = 1,526 |
| Nose only | Brand name only recorded, among children without an antibiotic | 1.3 (0.6 to 2.8); n = 615 | 0.9 (0.4 to 1.8); n = 716 |
| All cough and difficult breathing | Amoxicillin, among antibiotic-treated | 19.2 (16.6 to 22.1); n = 1,048 | 25.4 (22.6 to 28.4); n = 1,244 |
| All cough and difficult breathing | Cotrimoxazole, among antibiotic-treated | 3.6 (2.6 to 5.1); n = 1,048 | 5.3 (3.9 to 7.2); n = 1,244 |
| All cough and difficult breathing | Other oral antibiotic, among antibiotic-treated | 79.2 (76.3 to 81.9); n = 1,048 | 64.4 (61.0 to 67.6); n = 1,244 |
| All cough and difficult breathing | Injection or IV, among antibiotic-treated | 4.6 (3.4 to 6.2); n = 1,048 | 17.7 (15.0 to 20.7); n = 1,244 |
| All cough and difficult breathing | Two or more antibiotic types, among antibiotic-treated | 6.4 (4.7 to 8.6); n = 1,048 | 11.2 (9.3 to 13.5); n = 1,244 |
| All cough and difficult breathing | Amoxicillin, among all children in the group | 10.8 (9.2 to 12.5); n = 1,829 | 14.8 (13.1 to 16.7); n = 2,181 |
| All cough and difficult breathing | Brand name only recorded, among children without an antibiotic | 1.8 (1.0 to 3.2); n = 781 | 1.2 (0.6 to 2.4); n = 937 |

| Group | Pattern (mutually exclusive) | 2019 | 2025 |
| --- | --- | --- | --- |
| Suspected pneumonia | Any injection or IV | 9.6 (6.5 to 14.1); n = 303 | 19.2 (15.2 to 24.0); n = 429 |
| Suspected pneumonia | Amoxicillin only | 21.8 (16.7 to 27.9); n = 303 | 18.4 (14.6 to 22.9); n = 429 |
| Suspected pneumonia | Amoxicillin with another oral antibiotic | 3.4 (1.7 to 6.6); n = 303 | 5.1 (3.4 to 7.8); n = 429 |
| Suspected pneumonia | Cotrimoxazole without amoxicillin | 2.7 (1.3 to 5.6); n = 303 | 6.0 (3.9 to 9.2); n = 429 |
| Suspected pneumonia | Other oral antibiotic only | 62.5 (56.0 to 68.6); n = 303 | 51.2 (45.9 to 56.5); n = 429 |
| Nose only | Any injection or IV | 2.8 (1.8 to 4.3); n = 745 | 16.8 (13.3 to 20.9); n = 810 |
| Nose only | Amoxicillin only | 13.3 (10.7 to 16.4); n = 745 | 20.4 (17.3 to 23.9); n = 810 |
| Nose only | Amoxicillin with another oral antibiotic | 2.9 (1.6 to 5.1); n = 745 | 3.9 (2.4 to 6.1); n = 810 |
| Nose only | Cotrimoxazole without amoxicillin | 3.1 (2.0 to 4.9); n = 745 | 2.1 (1.2 to 3.6); n = 810 |
| Nose only | Other oral antibiotic only | 77.9 (74.4 to 81.0); n = 745 | 56.9 (52.5 to 61.2); n = 810 |

**Table S11. Where the antibiotic was obtained, by source of care, among treated children with cough and difficult breathing, 2019. Post hoc.**

Percentages (95% CI); several answers were allowed, so rows can exceed 100%. Informal or other includes traditional practitioners, shops, relatives, other places and don’t know.

| Source of care | n | Public health care | Private or NGO medical | Pharmacy | Informal or other |
| --- | --- | --- | --- | --- | --- |
| Public health care | 200 | 42.6 (35.1 to 50.4) | 2.3 (0.9 to 5.7) | 59.8 (52.0 to 67.2) | 9.2 (5.6 to 14.7) |
| Private or NGO medical | 276 | 1.5 (0.5 to 4.5) | 18.6 (14.1 to 24.1) | 81.7 (76.3 to 86.1) | 4.1 (2.2 to 7.3) |
| Pharmacy, no medical facility | 219 | 1.2 (0.3 to 3.9) | 2.1 (0.8 to 5.3) | 96.3 (92.7 to 98.2) | 2.7 (1.2 to 6.0) |
| Informal provider | 288 | 1.1 (0.3 to 4.0) | 2.5 (1.1 to 5.4) | 21.1 (16.2 to 26.9) | 77.1 (71.2 to 82.1) |
| No advice or treatment sought | 65 | 5.9 (2.3 to 14.3) | 0.0 | 63.0 (48.0 to 75.9) | 32.9 (20.5 to 48.2) |
| All treated children | 1,048 | 9.5 (7.7 to 11.6) | 6.4 (5.0 to 8.2) | 64.0 (60.7 to 67.2) | 25.3 (22.5 to 28.4) |
